# Vaccination and Variants: retrospective model for the evolution of Covid-19 in Italy

**DOI:** 10.1101/2022.02.27.22271593

**Authors:** Annalisa Fierro, Silvio Romano, Antonella Liccardo

## Abstract

The last year of Covid-19 pandemic has been characterized by the continuous chase between the vaccination campaign and the appearance of new variants that put further obstacles to the possibility of eradicating the virus and returning to normality in a short period. In the present paper we consider a deterministic compartmental model to discuss the evolution of the Covid-19 in Italy as a combined effect of vaccination campaign, new variant spreading, waning immunity and mobility restrictions. We analyze the role that different mechanisms, such as behavioral changes due to variable risk perception, variation of the population mobility, seasonal variability of the virus infectivity, and spreading of new variants have had in shaping the epidemiological curve. The fundamental impact of vaccines in drastically reducing the total increase in infections and deaths is also estimated. This work further underlines the crucial importance of vaccination and adoption of adequate individual protective measures in containing the pandemic.

## Introduction

The Covid-19 pandemic dramatically impacted on all aspects of world population life, causing a global lasting damage at economic, social and educational level and an enormous loss in terms of human lives. The extraordinary effort made worldwide for the development of vaccines against Covid-19 allowed an equally extraordinary achievement, such as the approval of the first vaccines in one year since the beginning of the pandemic. Such a circumstance made concrete the possibility of finally defeating the virus, limiting the further use of emergency measures, such as lock-downs, no longer economically sustainable.

The first vaccine authorized by the US Food and Drug Administration for distribution in the United States and by the European Medicine Agency (EMA) for the European Union (EU) countries were the mRNA-vaccine Comirnaty (BNT162b2), produced by BioNTech/Pfizer (December 2020) and soon after the one produced by Moderna (mRNA-1273). At the same time, the adenovirus viral vector vaccine Russian Sputnik V, produced by the Gamaleya Research Institute of Epidemiology and Microbiology, was authorized and distributed in different countries. One month later another adenovirus viral vector vaccine, the Vaxzevria (ChAdOx1-S), developed by the Oxford University and produced by Astrazeneca was authorized by EMA for distribution in EU. Afterwards, the single-dose Janssen COVID-19 vaccine was allowed to be distributed in the US and soon after (in March 2021) its distribution authorized also by EMA for the EU countries.

However, the vaccination campaign has suffered a series of setbacks, followed by successive accelerations, due to a number of circumstances. Firstly, the suspensions - and consequent restrictions of use [1, 2] - of the Astrazeneca and Janssen vaccines, due to rare cases of unusual blood clots with low blood platelets occurred in some vaccinated subjects [3], slowed down the vaccination campaign during the administration of the first doses in Italy. The summer holiday period was characterized by a significant reduction of the number of doses/per day administrated. It should be added that a communication not always clear and effective by the competent bodies, with the succession of different and sometimes antithetical recommendations of use for specific population age groups, also generated skepticism in a hard core of population that still shows reticence to undergo the vaccine. On the other hand, the gradual bureaucratic strengthening of the green pass that took place over the last few months has certainly led many previously reticent individuals to get vaccinated.

In Italy the vaccination campaign started in January 2021, firstly with sanitary personnel, and subsequently by age groups. At the time writing (31 January 2022) the percentage of Italian population fully vaccinated (one dose for Janssen vaccine and two doses for the other ones) is 77.4%, the percentage of those that received at least one dose is 84.3% and those with the booster dose 56.4%.

Effectiveness of the authorized vaccines has been estimated with different approaches [4, 5] either for mRNA vaccines [6–10] (with a range of estimated effectiveness between 91 and 95.3%), and for the adenovirus viral vector vaccine [11–13] (with effectiveness between 62, 1% and 90%), even if age specific effectiveness studies [14] show that the immunity peak response is lower in elderly people than in young population [15, 16]. The efficacy of the heterologous vaccine regimen has been discussed in [13] (estimated effectiveness 67% for heterologous ChAdOx1 nCoV-19 / BNT162b2 prime-boost vaccination, and 79% for heterologous ChAdOx1 nCoV-19 / mRNA-1273 prime-boost vaccination).

Nowadays there are two relevant aspects that significantly affect the success of the vaccination campaign against Covid-19 pandemic. The first one is the effectiveness of different vaccines on emerging variants of the virus [17, 18] and the second one is the mechanism of waning immunity, i.e. the decline of the vaccine efficacy as time passes [19, 20]. Both overlapping mechanisms lead to the potential occurrence of breakthrough infections among vaccinated individuals.

Most of the vaccines actually in use were developed against the virus wild-type and tested on large scale on Alpha variant (lineage B.1.1.7). However, the Delta variant (lineage B.1.617.2), first detected in India in late 2020, has spread worldwide, becoming soon the dominant strain in United Kingdom (approximately 90 − 95% cases from 7 to 21 June 2021) [21]. The European Centre for Disease Prevention and Control [22] confirmed the Delta variant to be dominant in the EU at the end of August (99, 6% prevalence with CI 72–100%). In Italy, according to the Italian National Institute of Health (ISS), the Delta variant increased in less than 5 months from 1% to 97.7% at the end of August.

Analysis by Public Health England [21] and by EPIcx lab in France [23] estimated the Delta variant to be at least 60% more transmissible than the Alpha variant and the vaccines to be less effective (after a single dose it was observed a 14% absolute reduction in vaccine effectiveness against symptomatic disease with Delta compared to Alpha, and a smaller 10% reduction in effectiveness after two doses [24]), whereas similar vaccine effectiveness against hospitalization was seen with the two variants. Also in Israel, Delta variant became dominant in July (with 90% prevalence) [25], with data on Pfizer vaccine effectiveness against hospitalization essentially in agreement with British data, but with a 30% absolute reduction in effectiveness against disease [26]. More optimistic values for the effectiveness of vaccination against symptomatic disease caused by the Delta variants are reported in [27], where it is evaluated at 88.0% for mRNA vaccine and to 67.0% for viral vector vaccine.

Both for the Comirnaty [28] and Moderna vaccine [29, 30], a booster dose with the same vaccine with the original SARS-CoV-2 spike protein has been strongly recommended to enforce the protection also against the Delta variant.

As stressed in [31] it is hard to differentiate the effectiveness reduction against new variant from the natural decay of immunity as time passes. According to the retrospective cohort study conducted in USA in the previously cited paper, the reduction in vaccine effectiveness against Covid-19 infections over time is probably primarily due to waning immunity with time rather than the Delta variant escaping vaccine protection.

In [33] the waning effectiveness of the vaccine in England, at 20 weeks or more after vaccination, was estimated to be 44.3% with Vaxzevria and 66.3% with Comirnaty, while the vaccine efficacy against hospitalization and death was confirmed. Similar results were found in data collected from the Israeli national database [34], and in Qatar [35]. It has been also ascertained that the immunity response decreases faster in elderly people than in young individuals [36].

This paper is devoted to the retrospective analysis of the evolution of the Covid-19 in Italy, keeping into account the vaccination rate, the variant spreading and the immunity decay. To this purpose we generalize the model developed in [37], with the introduction of the appropriate compartments for vaccinated individuals and suitably modify the parameters in order to simulate the increase in prevalence of the Delta variant, starting from mid-May and becoming dominant by August. Other variants, different from Alpha and Delta are not considered in the present model. The effect of waning immunity as time passes since the completion of the vaccine cycle and the occurrence of breakthrough infections is also included in the model.

We weigh the role that different mechanisms, such as variation in the population mobility, seasonal variations of the virus infectivity, variation in risk perception and appearance of new variants have had in forging the evolution of the epidemiological curve. We find that the most relevant mechanism is the seasonal variation in the stability of the virus, followed by the awareness mechanism, that induce individuals to increase/relax self-protective measures when the number of active cases increases/decreases. The appearance of the Delta variant and the mobility variations have had instead only marginal effects. We also estimate the effect of the vaccination campaign: in absence of vaccines the emerging scenario would have been dramatic with a percentage difference in the number of total infections and total deaths respectively equal to +63% and +55%. The model also predicts the appearance of a new variant (the Omicron variant) and its becoming dominant in January 2022.

## 1 The model

We use a compartment model similar to the one developed in [37] with the addition of appropriate vaccinated compartments. Furthermore, in order to follow the differences in the epidemic evolution among vaccinated (*v*) and unvaccinated (*u*) individuals, we split most of the compartments of the previous *SEI*_*A*_*I*_*S*_*I*_*D*_*RD* model in the vaccinated and unvaccinated sectors, indicated by the index *i*, with *i* = *v, u*. In particular individuals are divided in fifteen mutually exclusive classes according to their epidemiological status: the susceptible compartment *S*(*t*), the exposed vaccinated and unvaccinated compartments *E*^*i*^(*t*) (i.e. individuals that have been infected but are not yet infective), the vaccinated compartments *V*_*j*_ (with *j* = 1, 2, 3 indicating the first dose, the full vaccination, i.e. the second dose except for the single dose Janseen vaccine, and the third dose, respectively), the asymptomatic infected compartments 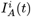, the symptomatic infective compartments 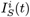, the diagnosed compartments 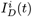, the dead compartments *D*^*i*^(*t*) and finally the recovered compartment *R*(*t*), that includes both unvaccinated and previously vaccinated individuals. We assume that healed individuals can loose immunity over time and return to the susceptible compartment, after an appropriate time. We do not consider demographic birth and (not related to the virus) death process. The epidemic dynamic is thus governed by the fluxes of individuals among these compartments, as shown in Fig.1, and it is fully described by a system of fifteen coupled first order differential equations for the normalized

**Fig 1.**
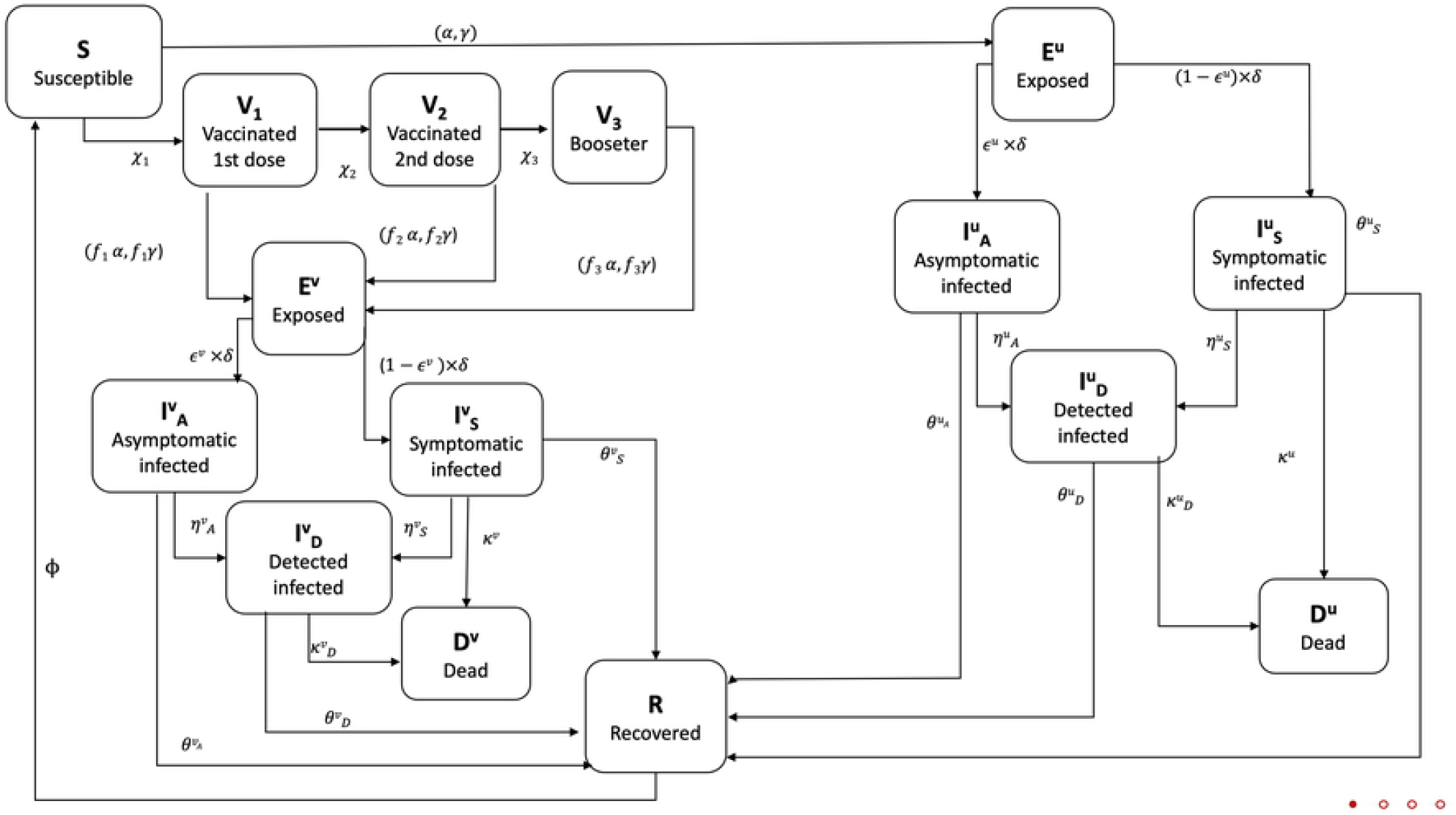
Flow chart summarizing the state variables, fluxes among compartments and related model parameters.

*SEV I*_*A*_*I*_*S*_*I*_*D*_*RDS* variables:

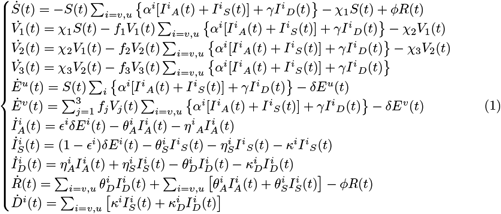

The system is closed and positive, i.e. all the state variables take non negative values for *t* ≥ 0, if initialized at time 0 with non negative values, and satisfy the mass conservation law 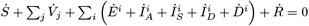, hence the sum of the states (the total population) is constant.

In our model individuals move to the vaccinated compartments *V*_*j*_ only when the vaccine protection becomes effective, two weeks after the inoculation.

Let us briefly review the main characteristic of the parameters. During the first wave of the pandemic, the world had to face with a completely novel virus and, as a consequence, it took some time to understand its mechanism of action, effective therapies and treatment modalities. As a consequence, most of the parameters that governed the evolution of the epidemic in the first period significantly changed in time. After more than one year, the knowledge and the experience in preventing, diagnosing and treating the infection made some of these parameters to reach an almost stable value. However some epidemiological parameters, as for instance individual mobility, seasonal stability of the virus, risk perception, immunity etc., are intrinsically time dependent. In order to fix them, we follow the same approach as in [37], i. e. we try to avoid step-functions and look for reasonable functional behaviors to describe their evolution. In the following we describe the principal time dependent parameters, while the constant ones are reported in Tables 3 and 6.

### The transmission rates, *α* and *γ*

The model has two different transmission rate parameters, *α* and *γ*, which govern the transmission of the virus respectively from undiagnosed and diagnosed individuals. The transmission from undiagnosed symptomatic or asymptomatic individuals is still the dominant mechanism in the spread of the epidemic. Following [38–40], we assume the transmission rates from undiagnosed vaccinated individuals, *α*^*v*^, to be different (and smaller) with respect to the corresponding rates for unvaccinated people, *α*^*u*^. Widely proven isolation protocols allow to assume the transmission of the virus by diagnosed cases, *γ*, to be residual and equally effective both from infected vaccinated and unvaccinated individuals, thus we do not differentiate the parameter *γ* between *u* and *v* individuals.

The parameters *α*^*i*^ can be both factorized in a pure contact term, *α*_*c*_, that we assume to be independent on the vaccination status^1^, describing the probability per unit time that a susceptible individual meets an infected individual, and the susceptibility term, *σ*^*i*^(*t*), which takes into account the probability that a potentially contagious contact between a susceptible and an infected individual leads to a new infection:

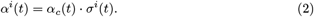

Both terms are in principle subject to changes. Restrictive measures, such as lock-down and limitations of access to places and services, certainly affect the contact rate, *α*_*c*_, as it happened during the first period of the pandemic. In [37] the global effect of such significant modifications of the mobility was encoded in an appropriate mobility function obtained as the weighted average on the mobility data from the Google Covid-19 Community Mobility Report [41]. Here we consider a different approach choosing to suitably modulate the contact term by assuming an increasing number of contacts in respect to February/March 2021, when Italy was still in lock down. In particular we choose an hyperbolic tangent transition function

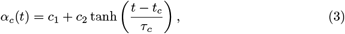

where the function is modulated in such a way to produce a doubling of the average number of contacts among individuals between February/March 2021 and the autumn period (i.e. *c*_1_ + *c*_2_ = 2 and *c*_1_ − *c*_2_ = 1), and *t*_*c*_ and *τ*_*c*_ are fitting parameters (the values of parameters are shown in Tables 4 and 5).

The susceptibility terms *σ*^*i*^(*t*) depend on different factors, partly related to the behaviors of individuals - e.g. more careful use of self-protective measures such as face masks, hand washing, due to increased risk perception during the rising phases of the epidemic - and partly related to the characteristics of the virus such as seasonal variation in the stability of the virus in airborne [42–44] and increased transmissibility due to the appearance of new Covid-19 variants. The functional form of the susceptibility function is thus chosen as

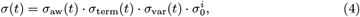

where *σ*_aw_ is the term encoding the awareness mechanism due to risk perception, *σ*_term_ encodes the modification of transmissibility due to seasonal variability, *σ*_var_ encodes the emergency of new variants and the last factor, 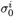, differentiate between vaccinated and unvaccinated individuals. Fixing 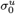 as a fitting parameter, we assume the transmissibility from vaccinated cases to be lower 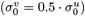, according to studies on household transmission [38, 45]. Recent literature seems to question the viral load of vaccinated infected individuals to be lower, however its decline seems to be faster than for unvaccinated individuals [46].

Following [37] we assume a prevalence based mechanism of rising awareness that increases the risk perception, inducing individuals to adopt more protective behaviors, with the effect of reducing the transmissibility of the virus during the peak. Thus we assume the awareness mechanism to act on the transmission rate by reducing it with a factor inversely proportional to the number of infective detected individuals, without any temporary effect of amplification or falsification [47]:

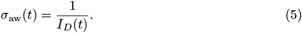

The reduction of contagiousness during the warm season, due to a potential decline of the stability of the virus in warm environment [42–45], already considered in [37], seems to be further confirmed by more recent literature [48–50]. Thus we mimic the susceptibility decrease/increase, in spring and autumn respectively, through appropriate hyperbolic tangent functions:

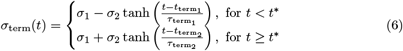

where 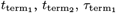 and 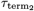 are fitting parameters ^2^. Finally the term

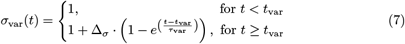

is the one encoding the susceptibility increase due to the circulation of new and more infective variants. In particular, following the ISS data [51] concerning the prevalence of Delta variant in Italy, the exponential increase is regulated in such a way to mimic the exponential transition from the susceptibility of the Alpha variant to the enhanced susceptibility of the Delta variant, that became dominant at the end of July (91.4% on July, 26). Following [21] and [23], Δ_*σ*_ is fixed a priori equal to 0.60.

Due to the circulation of the Delta variant also the parameter *γ* increases:

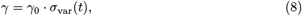

where *γ*_0_ is a fitting parameter.

### The susceptibility reduction function for vaccinated people, *f*_*j*_

Vaccinated people receive partial immunity against the infection, with an effectiveness that increases with the number of doses and decreases with the time occurred since the last inoculation and eventually with the appearing of new variants. In particular, the first dose gives only negligible immunity (estimated around 30% for both BNT162b2 and ChAdOx1 nCoV-19 vaccines in England [27]), whereas the third dose gives optimal immunization against the Delta variant [52] (estimated around 90% for BNT162b2 mRNA Vaccine in Israel). For what concerns the second dose, as previously discussed, different timing leads to extremely different levels of protection among the double dose vaccinated individuals. It has been nowadays established [53] that the effectiveness of vaccines against the infection decreases over a period of 6 months from the inoculation of the second dose (with an estimated decay of 40%), whereas the efficacy against severe manifestation is subject to a minor decay (less than 15%). This makes it difficult to quantify the average vaccine protection level to be included in an average field model such as the one considered in the present work. To this purpose we introduce a susceptibility reduction function for vaccinated people in Eq. (1) defined as *f*_*j*_ = 1 − *E*_*j*_, where *E*_*j*_ (with *j* = 1, 2, 3) is the vaccine efficacy after one dose, full vaccination and booster dose, respectively. Following literature, efficacy of first dose and booster dose are fixed equal to *E*_1_ = 30% and *E*_3_ = 90%, respectively. The evaluation of the mean efficacy over the fully vaccinated individuals, *E*_2_, is instead more complicated due to different timing of full vaccination.

The waning in time of different vaccines was evaluated for the US veterans in Ref. [53]. Using their data for the Pfizer-BioNTech vaccine (which is the most used in Italy in the first phase of the vaccination campaign) we find that these data are fitted with good approximation by the following function:

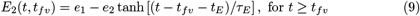

where *e*_1_, *e*_2_, *t*_*E*_ and *τ*_*E*_ are the parameters (shown in Tables 4 and 5), obtained fitting the data of Ref. [53], and *t*_*fv*_ is the timing of full vaccination (in [53], the month when full vaccination is administered; here instead *t*_*fv*_ is equal to 14 days after full vaccination).

The time-depending mean efficacy of full vaccination is then evaluated averaging over the fully vaccinated individuals (from which the number of individuals, who received the booster dose, was suitably subtracted) as follows:

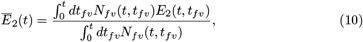

where *N*_*fv*_(*t, t*_*fv*_) are individuals that received full vaccination at (*t*_*fv*_ −14) days and have not received yet booster dose at (*t* −14) days.

Following [31] we assume the reduction in vaccine effectiveness against Covid-19 infections to be primarily due to the waning immunity mechanism, rather than the Delta variant escaping vaccine protection. Thus we do not consider a further reduction factor due to new variants.

Further parameters are listed below.

- *χ*_*j*_ (with *j* = 1, 2, 3), the vaccination rates corresponding to first, second (or single dose for the Janseen vaccine), and booster doses. As discussed in the introduction these parameters changed discontinuously during the last year, thus we fix them as step functions through the best fits of the experimental data. In Fig. 8 the simulated evolution and the time series of vaccinated individuals, as reported by ISS, are compared.
- *δ*, the inverse mean latent period assumed to be the same both for vaccinated and unvaccinated people. However, the emergency of new variants has been typically characterized by a reduction of the latent and incubation period [32]. Therefore we modulate its value according to the dominant variant:

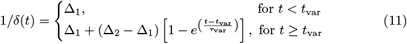

where Δ_1_ and Δ_2_ are fitting parameters, and *t*_var_ and *τ*_var_ are the same parameters present in Eq. (7).
- 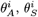 and 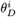, with *i* = *v, u*, the recovery rates respectively for asymptomatic, symptomatic, diagnosed vaccinated and unvaccinated individuals. The recovery rates for asymptomatic and symptomatic undiagnosed individuals are assumed not dependent on time. Those of diagnosed individuals have a significant variability over time, being affected by different factors, such as the test rate (increasing when the daily number of tests increases) and the number of active cases (decreasing when the sanitary system is overload). We assume 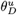 and 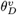 to be functions of time, not depending on the vaccination status of individual, i.e. 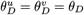, and following the form:

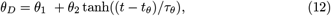

where *θ*_1_, *θ*_2_, *t*_*θ*_ and *τ*_*θ*_ are fitting parameters. This behavior corresponds to a decreasing in time of the number of days spent by infected diagnosed individuals in the diagnosed compartment, due to increased testing efficiency of the healthcare system through the involvement of analysis centers and pharmacies in testing operations.
- 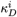 and *κ*^*i*^, with *i* = *v, u*, the mortality rates for diagnosed and undiagnosed infected individuals, respectively. The mortality rate for unvaccinated diagnosed individuals is assumed to follow the form:

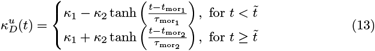

where *κ*_1_, *κ*_2_, 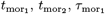 and 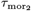 are fitting parameters ^3^. This behavior corresponds to a decreasing of the mortality during the summer period (from *κ*_1_ + *κ*_2_ to *κ*_1_ − *κ*_2_), probably due to the lower age of infected people, which returns to the same value of winter/spring in autumn. Following the data of ISS, the mortality rate for vaccinated diagnosed individuals is assumed to be lower than for unvaccinated people. In particular we assume the 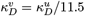, according to the relative risk estimation among vaccinated and unvaccinated individuals reported in [54] ^4^. Finally the mortality rate, *κ*^*i*^, for both vaccinated and unvaccinated undiagnosed individuals, is assumed equal to zero;
- *ϵ*^*i*^, with *i* = *v, u*, corresponding respectively to the asymptomatic percentages of infections among vaccinated and unvaccinated individuals. Although there is evidence that vaccinated individuals are protected from severe illness, to our knowledge, a systematic study of differences in the asymptomatic fraction of infection between vaccinated and unvaccinated individuals is still lacking. For this reason, we fix the fraction of asymptomatic vaccinated and unvaccinated individuals to be equal.
- 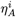 and 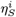, with *i* = *v, u*, corresponding respectively to the detection rates for asymptomatic and symptomatic infective individuals. These parameters are chosen to be constant in time and dependent on the vaccination status of individuals, both larger for unvaccinated than vaccinated people, reflecting the tendency of unvaccinated individuals to swab more easily than vaccinated ones.
- *ϕ*, corresponding to the rate at which recovered individuals loose immunity. On the time scales here considered, the presence of healed individuals, who return to being susceptible with a rate between 180^−1^ and 270^−1^ day^−1^ (as estimated in literature), turns out to be totally irrelevant, so we choose to set it equal to zero.

For the time dependence of the parameters in Eq.s (1) we preferred the use of hyperbolic tangents, except for Eq. (7), in which we adopted the exponential law - used also in Eq. (11) - obtained by fitting the real data for the prevalence of Delta variant in Italy. However, while in Eq. (9), the hyperbolic tangent comes up as a fit of the vaccine efficacy data, published in Ref. [53], in all other cases, the hyperbolic tangent was preferred because it allows to describe the crossover between two different values of the parameter by means of a continuous function, with derivatives of any order that are continuous in turn.

## 2 Results and discussion

The ODE Eqs. (1) have been solved using the SciPy libraries with initial conditions reported in Table 2 of Appendix. The best fit parameters, obtained minimizing the *χ*-square with respect to the experimental data, are listed in Tables 3, 4, 5 of Appendix.

Fig.s 2 compare the evolution of active detected cases (upper left), recovered detected cases (lower left) and dead detected cases (upper right) in our model, with the official data of the Italian outbreak reported daily by ISS, from February 20 to December 16, 2021, when the cases of Omicron variant, not included in the model, became not longer negligible. As we see in figures, through the introduction of some fundamental ingredients, the simulation manages to capture the trend of the real epidemic, not only qualitatively but also quantitatively.

**Fig 2.**
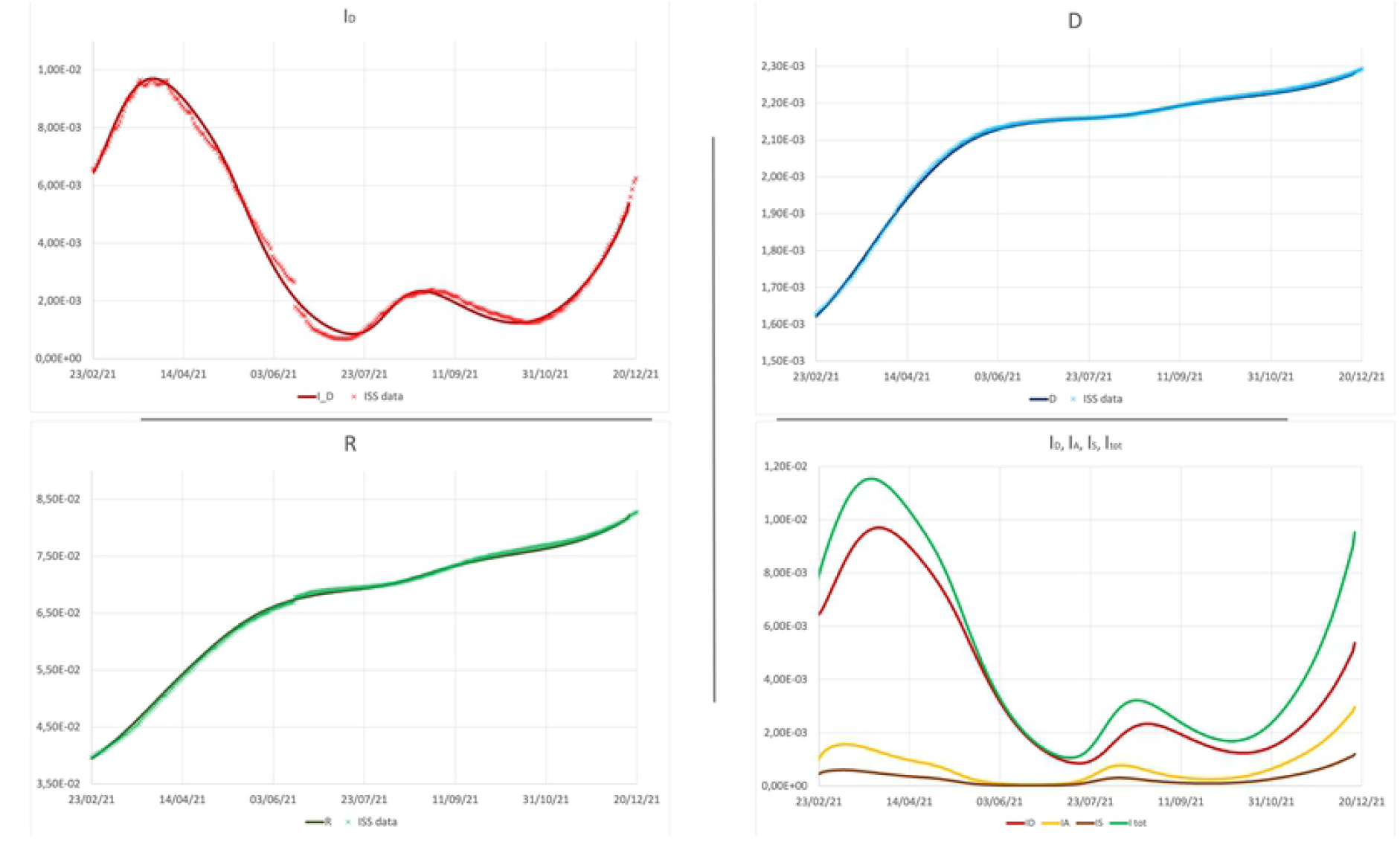
**Upper panel** - left: active detected cases; right: dead detected cases. **Lower panel** - left: recovered detected cases; right: detected, asymptomatic, symptomatic and total active cases.

Besides the detected cases, our model predicts a relevant number of undetected symptomatic and asymptomatic cases, as shown in Fig. 2 (lower right). According to our prevision, the percentage of undetected cases evolves from a minimum value of 3.6% at the beginning of summer, when the outbreak slowed down, to a value larger than 43% at the end of autumn. This circumstance can be related to the tendency within the large pool of vaccinated individuals to avoid testing when asymptomatic.

This mechanism is also useful to understand the result on the relative incidence among unvaccinated and vaccinated individuals. As shown in Fig. 3, our model predicts a different impact of the epidemic on vaccinated and unvaccinated individuals, in good agreement with data published by the ISS. In the inset of Fig. 3, the simulated relative incidence among unvaccinated detected cases and vaccinated detected ones (blue line in figure) is compared with the same quantity evaluated on the total cases (both detected and undetected) (green line), and with the experimental incidence (red dots), clearly evaluated only on detected cases. The blue line is typically slightly higher than the red dots, and systematically higher than the green line. This last circumstance is consistent with the hypothesis that, differently from unvaccinated individuals that are frequently required to test, vaccinated individuals, specially if asymptomatic, may remain undetected more frequently than unvaccinated ones.

**Fig 3.**
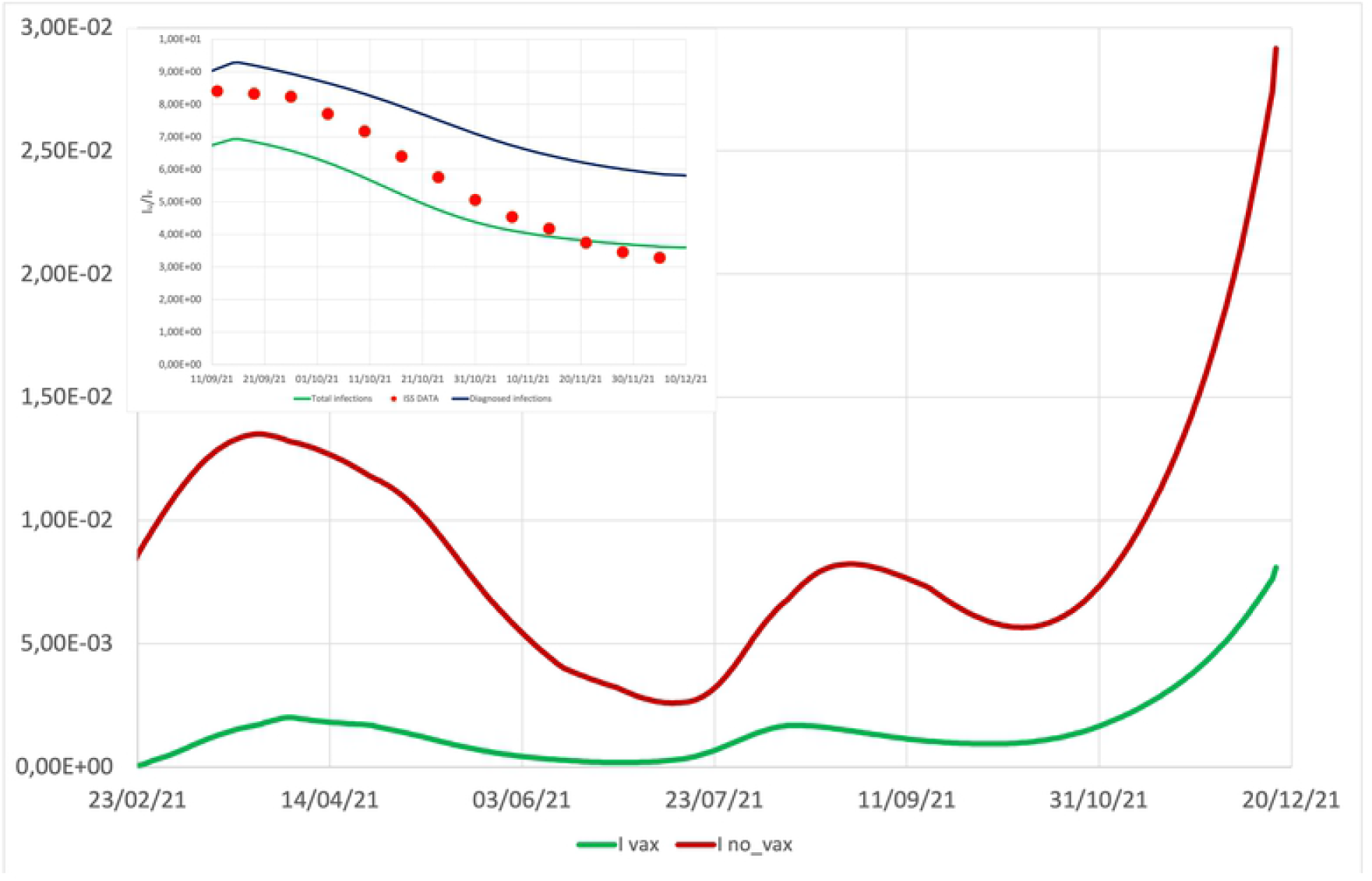
**Main**: incidence of total active cases (*I*_tot_ = *I*_*A*_ + *I*_*S*_ + *I*_*D*_) among vaccinated and unvaccinated individuals. **Inset**: the simulated relative incidence between unvaccinated detected cases and vaccinated detected ones, 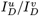 (blue line), is compared with the same quantity evaluated on both detected and undetected cases, 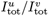 (green line), and with the experimental data (red dots), provided by the ISS.

When comparing model predictions and epidemiological data, it must be taken into account that both are not error-free. The error on real epidemiological data is difficult to assess, due to the stochasticity inherent in the epidemic spreading (an effect completely neglected in a deterministic compartmental model in a finite population with short-range interactions), but also due to the method of data acquisition that can introduce random, but also systematic, errors (see for instance the discontinuity present in the experimental data in mid-June due to an incorrect communication of the healed individuals by some Italian regions). A source of error is certainly linked to the diagnostic capacity, which has changed significantly during the epidemic and which is mainly linked to the amount of swabs that can be done daily.

Concerning the simulations, the main source of error is the estimation of epidemic control parameters, but also of the initial conditions. In order to reproduce the experimental curves, we had to fix many different parameters, concerning the efficacy of vaccine, the waning immunity mechanism, the spreading of new variants, and other parameters such as those appearing in Eq.s (7) and (9). It is reasonable to wonder how much the results obtained depend on the initial conditions and on these parameters set a priori. To explore these aspects, we carried out a sensitivity analysis focusing on one crucial number, such as the total number of cases diagnosed on the last day of the simulation, and evaluating how this number changes under modifications of the external parameters and initial conditions. We find that the results obtained are very stable for a fairly wide variation of the external parameters. In particular, Fig. 4 shows that the total number of detected cases at the final day of simulation (December 16, 2021) varies less than 1% by changing the vaccine efficacy parameters in Eq. (9) over a reasonable range of variability. Fig. 4 represents the total detected cases as a function of the maximum efficacy of full vaccination, *e*_1_ + *e*_2_, and the medium time of antibody decay, *t*_*E*_. It shows that the same total incidence is obtained when a decline of efficacy is counterbalanced by a suitable growth of the waning time. A similar variation less than 1% is observed by fixing *e*_1_ + *e*_2_ and changing the decay interval, *τ*_*E*_, in Eq. (9), together with *t*_*E*_ (data not shown).

**Fig 4.**
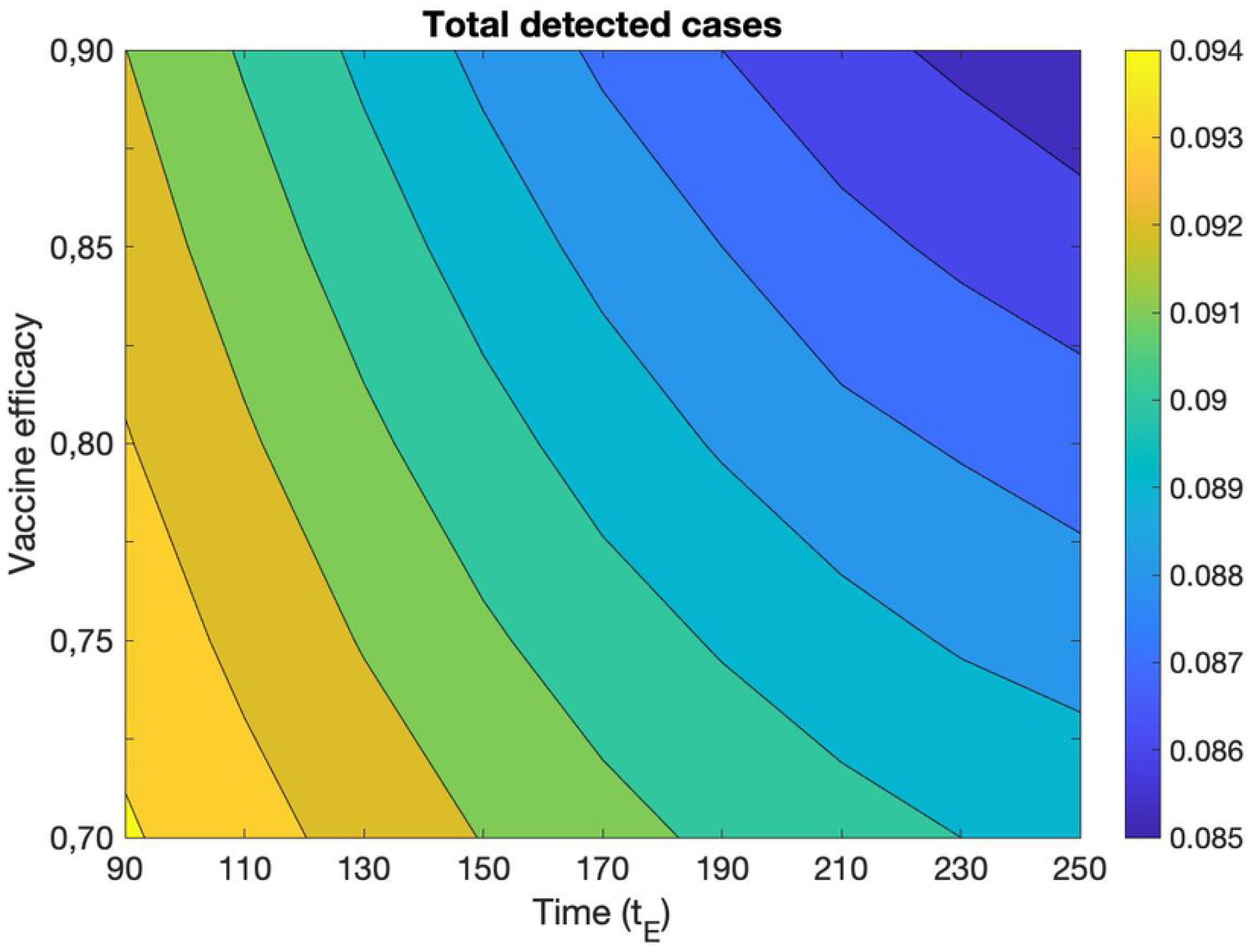
Total number of detected cases at the final day of simulation (December 16, 2021) as a function of the maximum efficacy of full vaccination, *e*_1_ + *e*_2_, and the medium time of antibody decay, *t*_*E*_.

Similarly it is interesting to explore what happens by varying the parameters regulating the insurgence of the Delta variant and the corresponding augmented transmissibility in Eq. (7). In particular by varying the increase in transmissibility, Δ_*σ*_, in the range [0.4, 1], the time of appearance of the new variant, *t*_*var*_, in the range [65, 105] and the time regulating the speed in the spreading of the new variant, *τ*_*var*_, in the range [10, 50] (Fig.s 5), the total number of detected cases at the final day of simulation varies less than 1%. Furthermore, it is observed that for values of Δ_*σ*_ small enough, it is completely irrelevant to vary *t*_*var*_ and *τ*_*var*_ in the established ranges. Similar findings are obtained varying the percentages of asymptomatic infected individuals, *ϵ*^*v*^ and *ϵ*^*u*^, or the initial conditions, in a reasonable range (data not shown).

**Fig 5.**
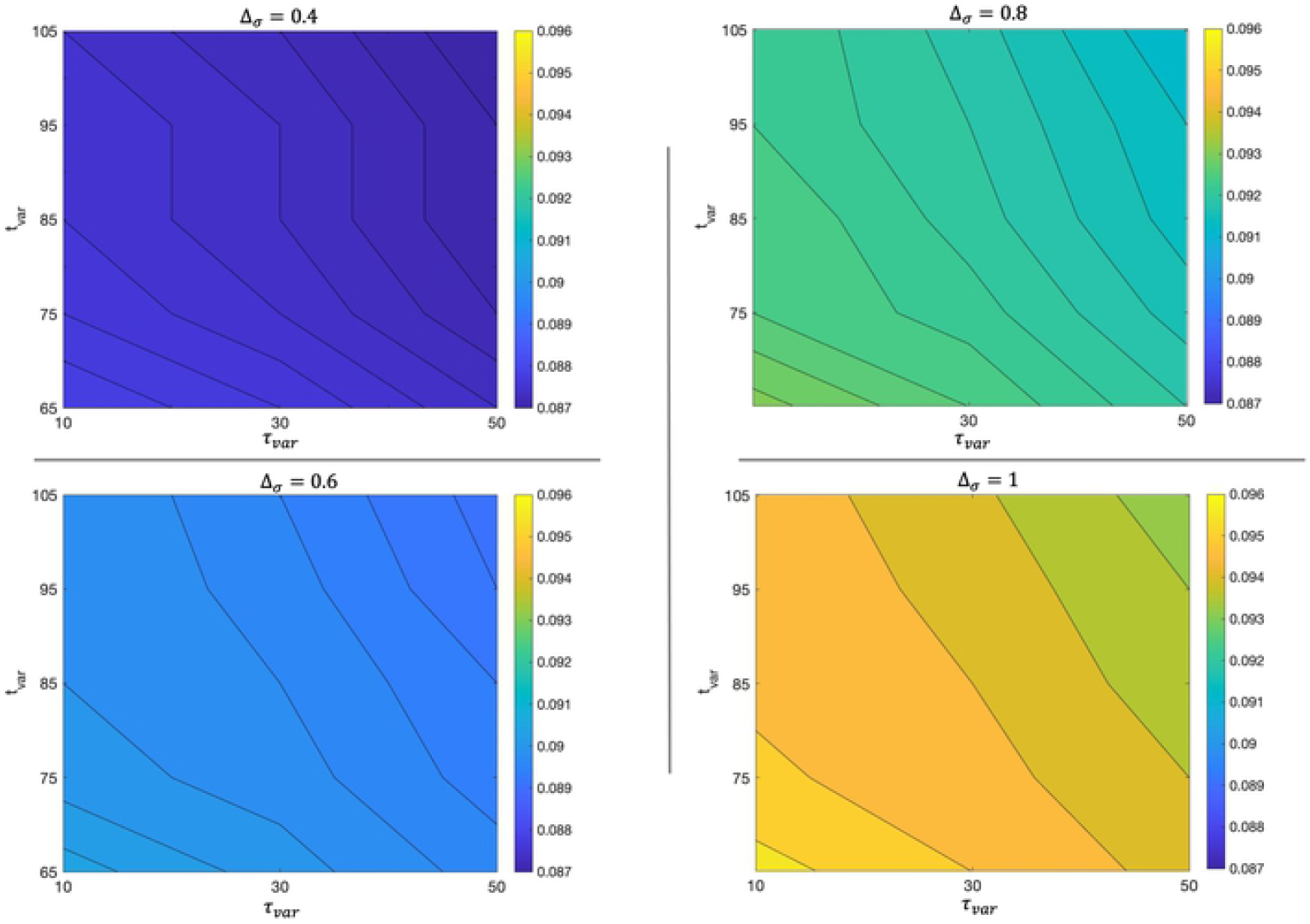
Total number of detected cases at the final day of simulation (December 16, 2021) for different values of the parameters Δ_*σ*_, *τ*_*var*_, *t*_*var*_.

In conclusion, the errors on the simulated data due to the estimation of initial conditions and of the parameters regulating efficacy of vaccine, waning immunity, spreading of the Delta variant and fraction of asymptomatic individuals, in realistic ranges of variation, do not seem to be greater than a few percent.

As previously seen, there are many different mechanisms that contribute to the evolution of the epidemic, such as the mobility of the population, the perceived risk, the seasonal variation in the virus stability, the appearance of new variants and, obviously, the vaccination campaign. All these mechanism are encoded in the model, through appropriate terms. It is thus interesting to analyze the contribution of each term to the epidemic evolution. Fig. 6 shows the trend of diagnosed cases in different scenarios obtained turning off one term at a time in Eq. (2) and (4) (and only for the *σ*_*var*_ also in Eq. (8)), or assuming the absence of vaccine. In particular, in Scenario I we disregard the effect of seasonal reduction of virus stability and infectivity during the summer period, in Scenario II we ‘freeze’ the awareness of individuals to the initial value, when the risk perception was quite high and the attention of individuals to respect sanitary measures, such as social distancing, frequent hand washing and wearing sanitary masks, was high as well. In Scenario III we consider what would have happened in absence of Delta variant. In Scenario IV we consider the effect of freezing the mobility to a low value, as the one during the lock-down period. Finally, Scenario V is devoted to understand what would have happened if there were no vaccines available.

**Fig 6.**
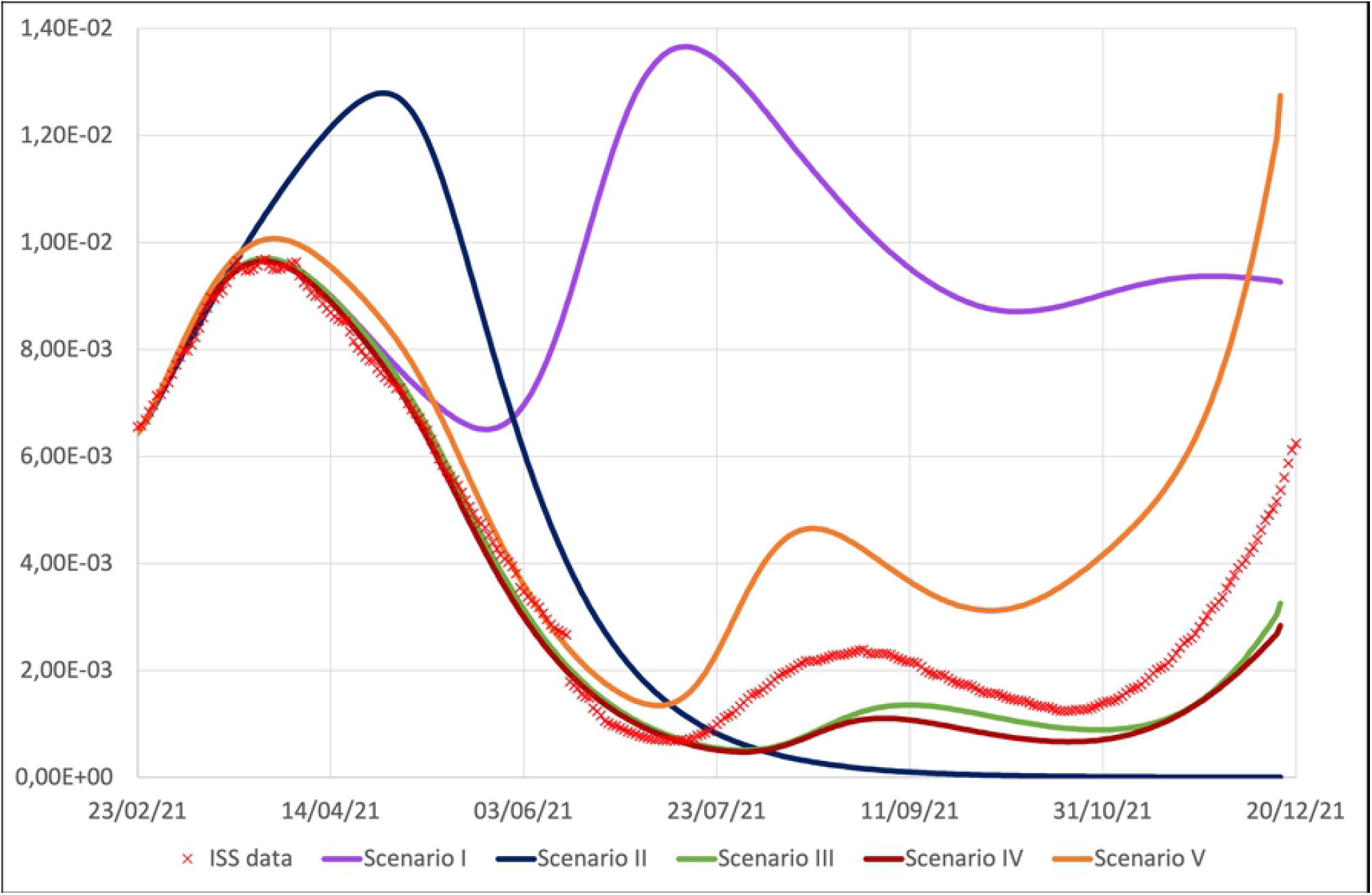
Diagnosed cases, *I*_*D*_, obtained turning off the seasonal term in Eq. (6) (Scenario I, violet line), the awareness term in Eq. (5) (Scenario II, blue line), the Delta variant term in Eq. (7) and (11) (Scenario III, green line), the mobility term in Eq. (3) (Scenario IV, brown line), and the vaccinations (Scenario V, orange line), compared with the ISS data.

For each scenario we evaluate the increase in the total infected detected cases and in total deaths with respect to the experimental initial values of ISS at time *t*_0_ 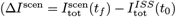 and Δ*D*^scen^ = *D*^scen^(*t*_*f*_) − *D*^ISS^(*t*_0_)) and in Table 1 we report the percentage variations of these quantities in the specific scenario with respect to those of the reference simulation ((Δ*I*^scen^ − Δ*I*^ref^)*/*Δ*I*^ref^ and analogously for the death term). As already observed in [37], we find that the seasonal modulation of virus transmissibility is an essential ingredient in order to explain the evolution of the epidemic curve during the summer period. Indeed by assuming this term to be constant (violet line in Fig. 6), and equal to its value in winter time, an agreement between data and simulation would have been obtained only in the first peak, then the curve for diagnosed cases, after the achievement of a minimum at the beginning of June, would start to rise again, reaching a second and significantly higher peak in summer, followed by a decrease to a long and still high plateau. In such a circumstance at the end of the simulation period (December 16, 2021) the number of total infections and total deaths would have been significantly higher (175.1% and 95.7% respectively) than the observed ones.

**Table 1.** Percentage variations in the number of total infected detected cases and total deaths with respect to the reference simulation at the final day of simulation (December 16, 2021).

| Scenario | % variation of $\Delta I_{\text{tot}}$ | % variation of $\Delta D$ |
| --- | --- | --- |
| I: no seasonal variations | +175.1% | +95.7% |
| II: no awareness mechanism | -12.4% | +8.5% |
| III: no Delta variant | -18.4% | -7.7% |
| IV: no mobility variations | -23.2% | -10.6% |
| V: no vaccines | +62.8% | +55.1% |

Analogously, if the awareness mechanism hadn’t been at work, a larger and higher peak would have been reached (blue line in Fig. 6), followed by a rapid decrease to zero of diagnosed individuals already around the beginning of September, so the subsequent increase in both the contact and seasonal terms would not have been sufficient to produce an increase in diagnosed cases. Let us note that this behavior corresponds to the situation in which the epidemic had spread always with the same high level of awareness that population had at the end of February, when Italy was in lock-down. But, in correspondence of the peak, the awareness is lower than in the reference simulation (due to the form of Eq. (5) and to the higher value of *I*_*D*_ in the peak), while it is larger in the summer period. As a consequence, this Scenario, is characterized by a negative percentage variation in Δ*I*_tot_ with respect to the reference simulation, while the percentage variation in Δ*D* is positive due to the enlargement and rise of the first peak, in a phase of epidemic with higher mortality.

Both contributions of the contact term, Eq. (3), and of the Delta variant, Eq. (7), have minor and very similar impacts on the epidemic evolution and their shutdown determines only the reduction of the summer peak and a slower autumn growth (brown and green lines in Fig. 6, respectively).

A separate analysis deserves the Scenario V with no vaccines. What would have happened in absence of vaccines? Fig. 6 shows the time evolution of active cases in absence of vaccinations (orange curve). During the first peak the curve does not differ much from the experimental one, consistently with the fact that the number of vaccinated individuals was still contained in this time window. However, from the summer period, and specially in autumn, when the vaccination campaign reaches a significant percentage of the population, scenario V foresees a significant increase compared to the case of the reference simulation/experimental data. In particular according to our simulation, if vaccines were not available, at the end of simulation, one would have obtained a percentage increase of 63% in the total detected infections and of 55% in the total deaths.

So far we have presented the model previsions up to mid-December. If we let the simulation run beyond this moment, the agreement between model predictions and real data becomes gradually worse. Actually this is not surprising because of the presence of the Omicron variant that became more and more important at the end of the year. On the other hand, the disagreement between the model and reality can give us a quantitative estimate of the presence of Omicron variant in Italy. In Fig. 7 the discrepancy between the diagnosed active cases provided by the ISS and the simulated data, *I*_*D*_, are plotted as a function of time. If we attribute the difference between observed and expected data to the presence of the Omicron variant, we can conclude that it appeared in Italy around the beginning of December, grew rapidly and became dominant in mid-January. This picture is in substantial agreement with the data provided by ISS [55], which gives prevalence estimates at the national level on January 3, 2022 for the Delta variant 19.22% (range: 0.0% − 66.7%) and for the Omicron variant 80.75% (range: 33.3% − 100%), despite the prevalence percentages are measured in a non-reproducible way within our model (the virus is sequenced by sample on positive swabs). The data in Fig. 7 may be indeed influenced by a different permanence of the individuals affected by the two variants of the virus in the compartment of the diagnosed individuals, for example if individuals with the Delta variant had on average a more severe disease, they would remain longer than the individuals affected by Omicron variant in the *I*_*D*_ compartment, systematically affecting the prevalence of the two variants estimated in this way.

**Fig 7.**
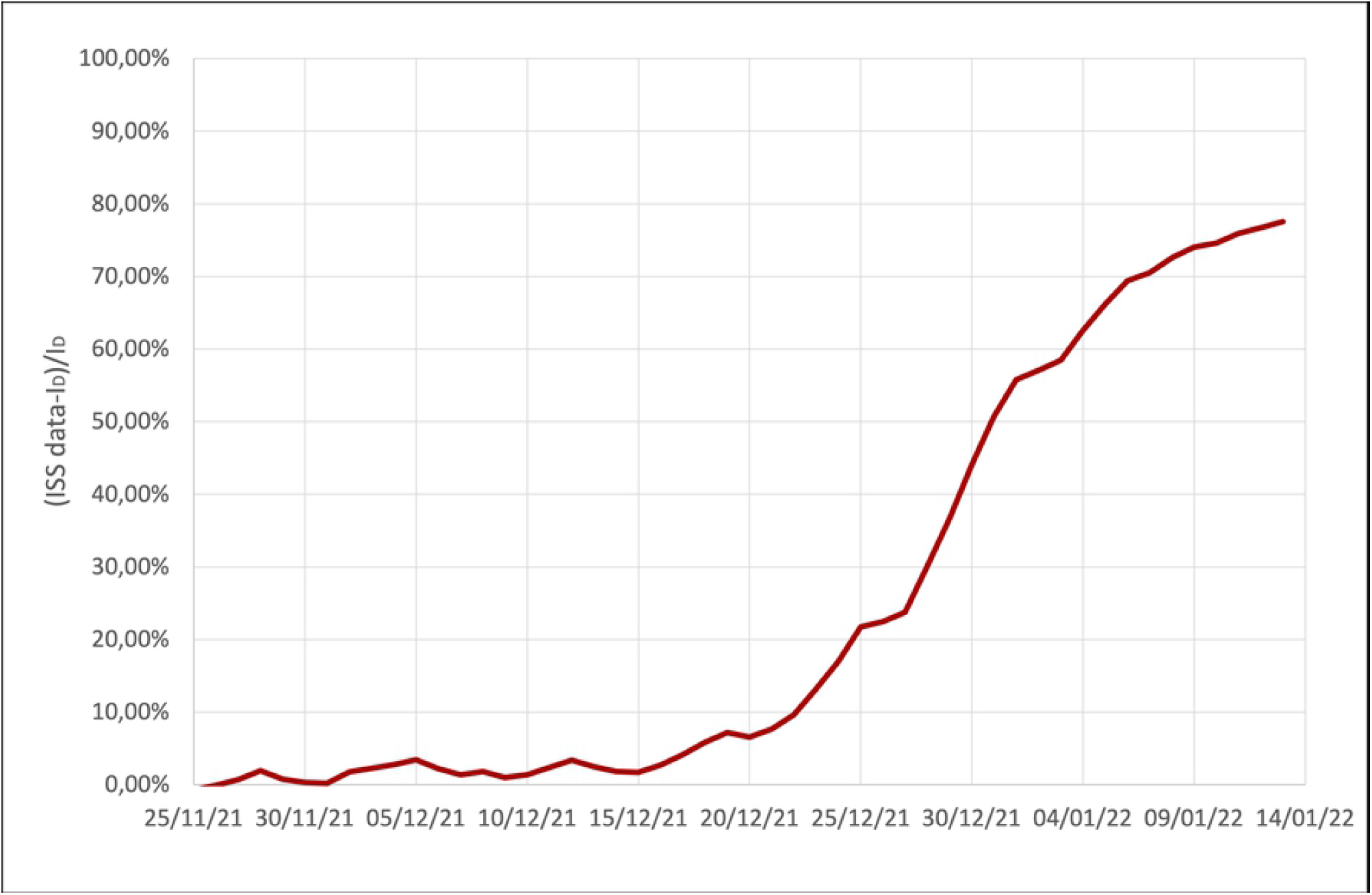
Discrepancy between the diagnosed active case provided by ISS and the simulated data, *I*_*D*_, during the insurgence of the Omicron variant.

The model has some limitations. The first one is that the system is assumed to be closed and protected from the injection of new cases from abroad. This circumstance is not fully justified, specially in the summer period, when the touristic flows increase. However, according to data published by ISTAT (National Institute of Statistics), even if the international tourist flow in 2021 was in recovery with respect to the year 2020 (+22, 3%), it was still far from the levels of 2019 (−38, 4%) [56]. The extension of the model to open system is left to future work.

Secondly, it has been shown that the vaccine efficacy to protect against severe infections is higher than the efficacy against mild or asymptomatic infection. Our model doesn’t distinguish the symptomatic cases according to the severity of symptoms, being pauci, mild and severe symptomatic cases, as well as hospitalized cases, all included in the symptomatic compartment. It would be interesting to rearrange the model in order to measure differences in hospitalization and severity of symptoms between vaccinated and unvaccinated individuals. However this would involve the introduction of new compartments into the model, circumstance that we avoided in order to keep the model simpler in the present paper.

In [57] authors study the attenuation of antibody titres after the second dose, showing that the most important factor in determining the waning immunity is sex, age and smoking. Our model does not take into account the age structure of population and not even the sex groups, and thus it is not able to capture differences in infections, mortality and recovering among different groups of individuals. We leave this interesting in-depth analysis to a future work.

Finally, it will be interesting to extend the stochastic models proposed in Ref.s [58, 59] for the pandemic H1N1 to the forth wave of Covid-19 epidemic: in this case indeed the almost total absence of mobility restrictions, which were instead present during the previous waves, would allow to apply the social contact hypothesis [60] in order to reproduce the epidemic spreading.

## 3 Conclusion

In conclusion, we presented a deterministic mean field model in which the vaccinated compartments with different number of doses have been suitably introduced. The model is able to reproduce the epidemic spreading in Italy during the third and in part the fourth wave of Covid-19. The analysis of the ingredients that must be taken into account in order to reproduce the epidemiological curves teaches a lot about the disease. The strong seasonal trend of the epidemic has been confirmed, together with the role of awareness mechanisms that allow to mitigate the epidemic spreading through individual protective behaviors adopted when the risk perception increases. The effects of the appearance of the Delta variant and the contact increase during the summer months were instead only marginal, causing a slight rise in the summer peak, being the mitigating effect of summer temperatures stronger. The model also predicts with remarkable accuracy the appearance of the Omicron variant and its becoming dominant in January 2022. According to our model, in absence of a vaccination campaign, the total number of infections and deaths would have been dramatically higher, confirming the fundamental role of the vaccine in containing the pandemic and saving human lives.

## Data Availability

All relevant data are within the manuscript and its Supporting Information files

## Appendix

The values of *R*_0_, *D*_0_, *V*_10_, *V*_20_, *V*_30_, reported in Table 2, correspond to the current values at the beginning of the period under examination, *t*_0_; since the number of vaccinated individuals is negligible at *t*_0_,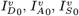 and 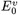 are put equal to zero; the initial values for 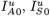 and 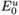, which are not experimentally observables, are obtained extrapolating simulation results from previous model developed in [37]; finally *S*_0_ is obtained as difference between 1 and the sum of the initial values of all the other compartments.

**Table 2.** Initial conditions (*N* is the Italian population, estimated 59258000 in 2021).

|  |  |
| --- | --- |
| $t_0$ | 19/02/2021 |
| $R_0$ | 2303199/$N$ |
| $D_0$ | 95235/$N$ |
| $V_{10}$ | 362101/$N$ |
| $V_{20}$ | 1085950/$N$ |
| $V_{30}$ | 0 |
| $I_{D0}^u$ | 382448/$N$ |
| $E_0^u$ | 364855/$N$ |
| $I_{A0}^u$ | 15297/$N$ |
| $I_{S0}^u$ | 7648/$N$ |

Fitting rates present in Eq.s (1), whose value is not dependent on time, are given in Table 3. Constants present in Eq.s (3–13), obtained as fitting parameters, are listed in Table 4. Days and intervals of time present in Eq.s (3–13), obtained as fitting parameters are reported in Table 5 (in the simulations the time 0 corresponds to the initial time, *t*_0_ reported in Table 2, i.e. February 19, 2021). Further parameters are reported in Table 6. Fig. 8 shows the comparison between the simulated evolution and the time series of vaccinated individuals, as reported by ISS.

**Table 3.** Fitting time independent rates in Eq.s (1).

|  |  |  |
| --- | --- | --- |
| $\theta_A^u$ | 1/7 day<sup>-1</sup> | recovery rate unvaccinated asymptomatic |
| $\theta_A^v$ | 1/7 day<sup>-1</sup> | recovery rate vaccinated asymptomatic |
| $\theta_S^u$ | 1/14 day<sup>-1</sup> | recovery rate unvaccinated symptomatic |
| $\theta_S^v$ | 1/10 day<sup>-1</sup> | recovery rate vaccinated symptomatic |
| $\eta_A^u$ | 1/7 day<sup>-1</sup> | detection rate unvaccinated asymptomatic |
| $\eta_A^v$ | 1/30 day<sup>-1</sup> | detection rate vaccinated asymptomatic |
| $\eta_S^u$ | 1/3 day<sup>-1</sup> | detection rate unvaccinated symptomatic |
| $\eta_S^v$ | 1/7 day<sup>-1</sup> | detection rate vaccinated symptomatic |

**Table 4.** Fitting parameters in Eq.s (3–13).

|  |  |  |
| --- | --- | --- |
| $c_1$ | 1.5 | Eq.(3) |
| $c_2$ | 0.5 | Eq.(3) |
| $\sigma_0^u$ | 0.00280 day<sup>-1</sup> | Eq.(4) |
| $\sigma_1$ | 0.57 | Eq.(6) |
| $\sigma_2$ | 0.43 | Eq.(6) |
| $\gamma_0$ | 0.001 day<sup>-1</sup> | Eq.(8) |
| $e_1$ | 0.6 | Eq.(9) |
| $e_2$ | 0.3 | Eq.(9) |
| $\Delta_1$ | 8 days | Eq.(11) |
| $\Delta_2$ | 5 days | Eq.(11) |
| $\theta_1$ | 0.0375 day<sup>-1</sup> | Eq.(12) |
| $\theta_2$ | 0.0045 day<sup>-1</sup> | Eq.(12) |
| $\kappa_1$ | 0.00055 day<sup>-1</sup> | Eq.(13) |
| $\kappa_2$ | 0.00021 day<sup>-1</sup> | Eq.(13) |

**Table 5.** Time intervals and reference days in Eq.s (3–13), obtained as fitting parameters.

|  |  |  |  |  |  |
| --- | --- | --- | --- | --- | --- |
| $t_{\text{term}_1}$ | 70 | 30/04/2021 | $\tau_{\text{term}_1}$ | 10 days | Eq.(6) |
| $t_\theta$ | 75 | 05/05/2021 | $\tau_\theta$ | 30 days | Eq.(12) |
| $t_{\text{mor}_1}$ | 85 | 15/05/2021 | $\tau_{\text{mor}_1}$ | 40 days | Eq.(13) |
| $t_{\text{var}}$ | 85 | 15/05/2021 | $\tau_{\text{var}}$ | 30 days | Eq.(7,11) |
| $t_c$ | 120 | 19/06/2021 | $\tau_c$ | 40 days | Eq.(3) |
| $t_E$ | 130 | 29/06/2021 | $\tau_E$ | 90 days | Eq.(9) |
| $t_{\text{mor}_2}$ | 200 | 07/09/2021 | $\tau_{\text{mor}_2}$ | 40 days | Eq.(13) |
| $t_{\text{term}_2}$ | 295 | 11/12/2021 | $\tau_{\text{term}_2}$ | 30 days | Eq.(6) |

**Table 6.** Further parameters, whose values are fixed a priori.

|  |  |  |
| --- | --- | --- |
| $\Delta_\sigma$ | 0.60 | transmissibility rise due to Delta variant, Eq.(7) |
| $\epsilon^v = \epsilon^u$ | 0.65 | asymptomatic infected individual percentage |
| $f_1$ | 0.70 | susceptibility reduction function after one vaccine dose |
| $f_3$ | 0.10 | susceptibility reduction function after booster |
| $\phi$ | 0 day<sup>-1</sup> | immunity lost rate |

**Fig 8.**
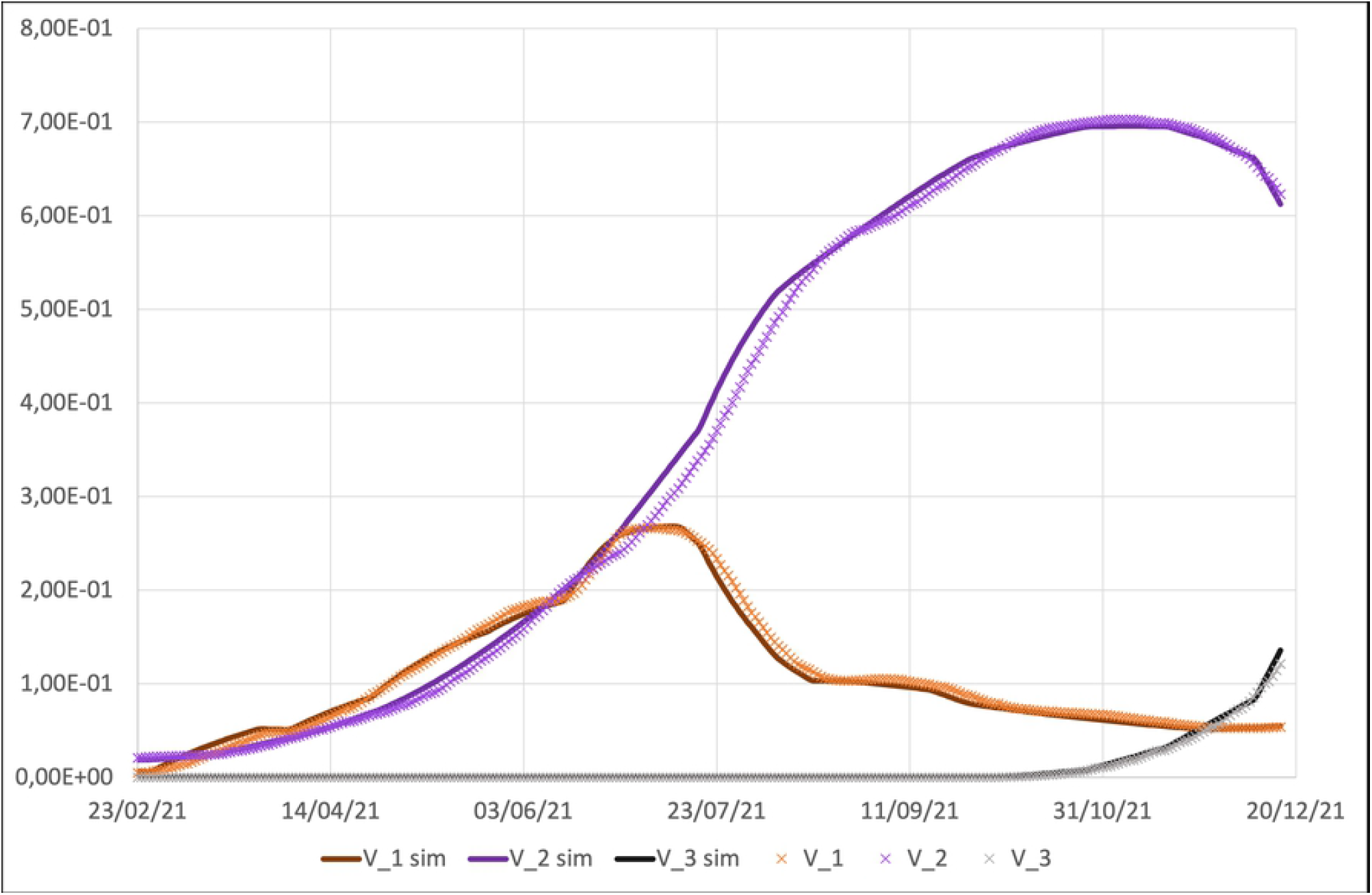
Best fit of vaccinated individuals. *χ*_*j*_ (*j* = 1, 2, 3) in Eq.s (1) are step functions suitably chosen to reproduce real data.

As already stressed in [37] most of the epidemiological compartmental models published in literature include more than three compartments and a relevant number of parameters that regulate fluxes between these compartments, within the specific model. Few available observables (i.e. data sets) do not allow to univocally fix the parameter values in the phase space. The basic idea at the base of such modelization is to exhibit a possible set of parameters, that, within the specific model/formulation, allows to reproduce the epidemic evolution.

## Acknowledgments

A.L. and A.F. acknowledge financial support of the MIUR PRIN 2017WZFTZP “Stochastic forecasting in complex systems”. The funders had no role in study design, data collection and analysis, decision to publish, or preparation of the manuscript.

## Footnotes

1 During the period under investigation, selective mobility restrictions for unvaccinated individuals were not yet at work.

2 *t*\* is any time ≫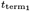 and ≪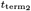, such that the two hyperbolic tangent functions assume the same asymptotic value. For simplicity we put 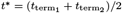.

3 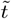 is any time ≫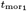 and ≪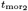, such that the two hyperbolic tangent functions assume the same asymptotic value. For simplicity we can put 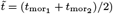.

4 According to [54], the relative mortality risk between vaccinated and unvaccinated individuals is 9.0, 11.5 and 30.3, respectively, for individuals fully vaccinated more than 120 days before, for those fully vaccinated less then 120 days before and for those with booster dose. During the period under investigation, the number of individuals with booster dose was negligible and mostly of the population recently completed the vaccination cycle.

